# Genetic Polymorphisms, Adherence to Mediterranean Diet and Microbiota-Associated Urolithin Metabotypes: A Complex Cocktail to Predict the Obesity in Childhood-Adolescence

**DOI:** 10.1101/2019.12.11.19014431

**Authors:** Adrián Cortés-Martín, Gonzalo Colmenarejo, María Victoria Selma, Juan Carlos Espín

**Affiliations:** Laboratory of Food & Health, Research Group on Quality, Safety and Bioactivity of Plant Foods, Dept. Food Science and Technology, CEBAS-CSIC, P.O. Box 164, 30100 Campus de Espinardo, Murcia, Spain; Biostatistics and Bioinformatics Unit, IMDEA Food Institute, Crta. de Cantoblanco nº 8, 28049 Madrid, Spain

## Abstract

Environmental and genetic factors are associated with pandemic obesity since childhood. However, the association of overweight-obesity with these factors, acting as a consortium, has been scarcely studied in children. We aimed here to assess the probabilities of being overweighed-obese in a randomly recruited cohort of Spanish children and adolescents (*n*=415, 5–17 years-old) by estimating the odds ratios for different predictor variables, and their relative importance in the prediction. The predictor variables were ethnicity, age, sex, adherence to the Mediterranean diet (KIDMED), physical activity, urolithin metabotypes (UM-A, UM-B and UM-0) as biomarkers of the gut microbiota, and 53 single-nucleotide polymorphisms (SNPs) from 43 genes mainly related to obesity and cardiometabolic diseases. A proportional-odds logistic ordinal regression, validated through bootstrap, was used to model the data. While every variable was not independently associated with overweight-obesity, however, the ordinal logistic model revealed that overweight-obesity prevalence was related to being a young boy with either UM-B or UM-0, low KIDMED score and high contribution of a consortium of 24 SNPs, being rs1801253-*ADRB1*, rs4343-*ACE*, rs8061518-*FTO*, rs1130864-*CRP*, rs659366-*UCP2*, rs6131-*SELP*, rs12535708-*LEP*, rs1501299-*ADIPOQ*, rs708272-*CETP* and rs2241766-*ADIPOQ* the top-ten contributing SNPs. Additional research should confirm and complete this model by including dietary interventions and the individuals’ gut microbiota composition.

## Introduction

Obesity is an aetiological condition associated with some types of cancer and cardiometabolic diseases such as type-2 diabetes, metabolic syndrome, non-alcoholic steatohepatitis, and hypertension^1^. Nowadays, it is widely accepted that the combination of high-energy diets, genetic make-up, sedentary lifestyles and gut dysbiosis (impaired composition and functionality of the gut microbiota) are involved in the obesity pandemic^2,3,4^. The prevalence of these significant health threats has risen to shocking proportions worldwide, including countries like Spain with ancestral adherence to the Mediterranean diet^5^. The increase in the rate of obesity has been mainly attributed to the ‘Westernization’ of the diet, the decrease of physical activity from childhood, currently aggravated by the abuse of playing videogames, and the increasingly early access to digital devices such as smartphones^6^. However, other potential variables could be participating in this pandemic.

Many studies have associated obesity in children and adults with single nucleotide polymorphisms (SNPs). For example, in genome-wide association studies (GWAS), the rs9939609 SNP in the fat-mass-and-obesity-associated (*FTO*) gene has been reported to account for a modest, but a statistically significant, increase of 0.4 kg/m^2^ body mass index (BMI) units for each risk allele (A)^7^. Although it is not fully understood yet, this association is partially mediated via controlling feeding behaviour^8^. However, not all the studies report this association, including those conducted in the child population^9,10^. Indeed, more than one million SNPs have been detected in the human genome^11^, and thus, instead of only one or few specific SNPs, it is more conceivable to expect the complex action of a consortium of SNPs potentially interacting with many other variables and associated with different conditions, including obesity.

The gut microbiota is also involved in the pathophysiology of obesity, although the associated mechanisms are not fully known yet^3^. Nevertheless, a number of pathways have been identified such as the translocation of lipopolysaccharides (LPS) from the gut to the bloodstream^12^, the regulation of gut hormones, energy harvest, inflammatory responses, lipogenesis and immune interactions^13^ as well as the regulation of white adipose tissue inflammation via microRNAs^14^. Recently, a potential nexus between the dissimilar metabolism of some dietary constituents by the microbiota and obesity has been proposed. This has been suggested for the metabolism of the polyphenols isoflavones^15^ and ellagitannins^16,17,18^ that yield specific metabolites, that is, equol and(or) *O*-desmethylangolensin (ODMA) in the case of isoflavones^19^, and different urolithin combinations in the case of ellagitannins^17^. These particular metabolisms give rise to specific metabolizing phenotypes (so-called ‘metabotypes’^16^) such as equol and(or) ODMA ‘producers’ vs ‘non-producers’ in the case of isoflavones^19^, and also the urolithin metabotypes associated with the metabolism of ellagitannins, i.e., metabotype A (UM-A; individuals that produce only urolithin A), B (UM-B; production of isourolithin A, urolithin B and also urolithin A) and 0 (UM-0; urolithin non-producers)^17^. The occurrence of specific gut microbiota metabotypes is behind the inter-individual variability upon polyphenol consumption^20,21^ and could be indirect markers of gut dysbiosis reflecting the individuals’ gut microbiota composition, richness, diversity, and functionality^22,23,24^. Although the gut microbiota associated with UM-B and UM-0 individuals show a dysbiotic-prone pattern^24^, however, the unequivocal association between these metabotypes with obesity has not been confirmed so far due to its multifactorial aetiology^24,25^.

In the present study, we aimed to assess the probabilities of being overweighed or obese in a cohort of children and adolescents from the Southeast of Spain by estimating the odds ratios (ORs) for different predictor variables and their relative importance in the prediction of the response. In this proof-of-concept, we considered as predictor variables the urolithin metabotypes as biomarkers of the gut microbiota, ethnicity, age, sex, the adherence to the Mediterranean diet, physical activity, and a consortium of 53 SNPs from 43 genes mainly related to obesity and cardiometabolic diseases.

## Methods

### Study Population

This research (‘*The PolyMicroBio study’*) was included in the Spanish National Project AGL2015-64124-R and complied with the ethical guidelines outlined in the Declaration of Helsinki and ethical principles for medical research involving human subjects (Seoul, Korea, 2008). The study was conceived to stratify the participants according to their urolithin metabotypes after three days of walnuts or pomegranate juice consumption^25^ and was not intended to modify any variable in the children. The trial was registered at clinicaltrials.gov (NCT03318042), and the Spanish National Research Council’s Bioethics Committee (Madrid, Spain) approved the protocol. Inclusion criteria were ages from 5 to 17 years old and good health status. Exclusion criteria were diagnosed pathology, previous gastrointestinal surgery, chronic medication and antibiotic intake one month before participating. A total of 415 children and adolescents were randomly recruited. Children within the 5 to 12 years old group (*n* = 202) were recruited from the public primary school ‘CEIP Jara Carrillo’ (Alcantarilla, Murcia, Spain) and adolescents aged from 13 to 17 (*n* = 213) from the public high school ‘IES Alcántara’ (Alcantarilla, Murcia, Spain). Parents were fully informed and gave their written informed consent before the participation of all students.

### Urolithin metabotypes

Children and adolescents consumed 25 g peeled raw walnuts daily or 250 mL of pomegranate juice daily (in the case of individuals allergic to nuts) for three days. Packs of peeled walnuts were kindly provided by Borges International Group, S.L. (Reus, Tarragona, Spain) and pomegranate juice by the AMC Group (Espinardo, Murcia, Spain). In the morning of the fourth day, a sample of urine was provided for its analysis by high-performance liquid chromatography with diode array detection coupled to electrospray ionisation and ion-trap tandem mass spectrometry (HPLC-DAD-ESI-IT-MS/MS), and ultra-high performance liquid chromatography coupled with electrospray ionization-quadrupole-time-of-flight-mass spectrometry (UPLC-ESI-QTOF-MS) as described elsewhere^26^. This allowed the stratification of the participants according to their different capacity to metabolise ellagic acid derivatives into urolithins, i.e., urolithin metabotypes UM-A, UM-B or UM-0 as previously described^25^.

### Anthropometric measurements and validated questionnaires

The determinations of height, weight, and waist and hip circumference were performed always by the same research staff, using the same equipment in all cases, and in the presence of teachers from the educational centres. The child growth standards from the World Health Organization (WHO) were used to define the BMI (kg/m^2^) cut-offs for underweight, normoweight, overweight and obese individuals as a function of sex and age^5^. The students were asked to record possible incidences (medication, protocol compliance, etc.), and also their physical activity level^27^, which took into account the two hours of physical activity a week in their schools (low activity) and the practice of additional extracurricular sports at least three days a week (high activity). Besides, a validated questionnaire to assess the adherence to the Mediterranean diet in children (KIDMED) was used^28^. The score in this questionnaire (ranging from 1 to 13, from very poor to optimum adhesion, respectively) was grouped as ‘Low’ (score from 1 to 4), ‘Medium’ (from 5 to 8), and ‘Good’ (from 9 to 13).

### Selection of SNPs and genotyping

Candidate genes and polymorphisms were identified after browsing the Single Nucleotide Polymorphism Database (dbSNP) and examining the published literature regarding each known gene and variant (favourable and unfavourable) associations^29,30,31^. On the same day of the anthropometric evaluation, saliva samples were obtained by gently rubbing the inside part of the cheek with a sterile swab, free of human RNA and DNA (Deltalab, Barcelona, Spain). Children were asked to clean their mouths and avoid eating or drinking 60 min before collection of samples to prevent contaminations. Two samples were obtained per student. The swabs were immediately stored in refrigeration and further frozen at −80 °C until their processing. Genomic DNA extraction and genotyping were carried out at the GENYAL Platform (IMDEA-Food, Madrid, Spain) using the OpenArray™ AccuFill™ System (Life Technologies Inc. Carlsbad, CA, USA) as described elsewhere^32^. Data analysis was made by TaqMan Genotyper Software v1.3 (autocaller confidence level > 90%).

### Statistical analysis

A proportional-odds logistic ordinal regression was used to model the data with the software R version 3.5.1 (www.r-project.org). Nine subjects with > 40% missing SNPs were removed, resulting in a final sample size of 406 students. The missing data was singly imputed using the missForest R package. Redundant predictors (rs9928094-*FTO*, rs9935401-*FTO*) were identified and removed using the Hmisc R package. SNPs with either favourable or unfavourable genotype frequencies below 5% were also removed (rs4994-*ADRB3*, rs7913948-*ALOX5*, rs7412-*APOE*, rs328-*LPL*, rs16139-*NPY*, rs6008259-*PPARγ*, rs2066826-*PTGS2*) (Supplementary Table 1). Using the ‘n/15 rule’^33^, only 21 predictors could be used to get reliable estimates. Consequently, data reduction was applied to the SNPs by applying Multiple Correspondence Analysis (MCA) and using only the first 15 MCA dimensions. Ethnic groups representing less than 1% each were merged into the ‘Other’ category.

Wald tests for all the predictors in the model were generated, and they were further ranked by importance based on the χ^2^-degrees of freedom (-df) score. A simplified model was obtained by applying a ‘fast-backwards’ variable elimination approach^34^ based on the Akaike’s Information Criterion (AIC)^35^. The approximate βs and ORs (and their 95% confidence intervals, CI) of the remaining variables were reported. The full model was validated through bootstrap to provide estimates of the performance of the model in new data in comparison with the training data, in the form of Sommers D_xy_, R^2^, intercept, slope, E_max_, and Briers B score. The significant contributions (coefficients of determination R^2^ with *p* values < 0.05) of SNPs to the essential MCA dimension were plotted to deconvolute it.

We used the proportional odds assumption in the model. ‘Physical activity’, ‘KIDMED’ and ‘Urolithin metabotype’ showed some deviation from this assumption. However, alternative extended continuation ratio models with this assumption relaxed for these variables did not result in better models according to the AIC, i.e., the higher complexity of the model was not compensated by the increase in the fit. The inclusion of transformations of some predictors, including a restricted cubic spline for both ‘Age’ and ‘KIDMED’, resulted in improved models, as judged by the AIC. Finally, a genetic score (computed as the sum of risk alleles) was also tested as a possible surrogate for the SNPs variables, but this did not result in a better replacement for the MCA dimensions. All tests were bilateral, with a significance level of 0.05.

Other statistical analyses were carried out using the SPSS software, v23.0 (SPSS Inc., Chicago, IL, USA). When more than two groups were compared, analyses of variance (ANOVA), followed by Bonferroni-corrected *t*-test (for post-hoc analysis) or the Kruskal–Wallis followed by Dunn’s test were used for normally and non-normally distributed data, respectively (KIDMED score vs *FTO* genotype TT, AT or AA, etc.). Comparison of non-normally distributed quantitative variables between two clusters was approached using the Mann-Whitney U-test (*FTO* TT genotype vs BMI or waist, etc.). Comparison of categorical variables was assessed using the Pearson’s χ ^2^ test. Spearman’s rank or Pearson correlations were applied to explore possible associations between variables (BMI vs hip/height, etc.). Plots of data were performed using Sigma Plot 13.0 (Systat Software, San Jose, CA, USA).

## Results

### Characteristics of the Cohort and Associations with Overweight-Obesity

Table 1 shows characteristics of the cohort as a function of age and sex, including anthropometric values (hip, waist, weight and BMI), the distribution of urolithin metabotypes (A, B and 0), the KIDMED scores (grouped as low, medium and good adherence), physical activity and the percentage of normoweight, overweight and obesity. The participants were mainly Caucasian-Europeans (93.5%), with a small proportion of Arabs (2.9%) and Amerindians (2.2%), and a marginal presence of Black-Africans (0.96%), Asian-Chinese (0.22%), and Indo-Aryans (0.22%).

**Table 1.** Characteristics of the study population. *Results are expressed as median and (range); F, Female; M, Male; KIDMED (L/A/G): Low quality, Medium quality and Good quality diet, respectively. NW, normoweight; OW, overweight; OB, obesity; Physical activity: H, high; L, low.

| Age (y) | Sex | Hip (cm)* | Waist (cm)* | Weight (kg)* | Height (cm)* | BMI (kg/m<sup>2</sup>)* | NW (%) | OW (%) | OB (%) | KIDMED (%)<br>(L/M/G) | Physical Activity<br>(%) (H/L) | Metabotype (%)<br>(A/B/O) |
| --- | --- | --- | --- | --- | --- | --- | --- | --- | --- | --- | --- | --- |
| 5 | F (n=16) | 65.5 (58.0-81.0) | 58.5 (48.0-76.0) | 20.9 (15.9-31.1) | 110.5 (103.5-119.0) | 16.4 (13.6-25.7) | 50.0 | 18.8 | 31.2 | 28.6/64.3/7.1 | 76.9/23.1 | 81.3/0.0/18.7 |
|  | M (n=14) | 66.5 (58.0-83.0) | 59.5 (51.0-71.5) | 22.2 (17.4-34.2) | 114.5 (110.0-124.0) | 16.7 (13.7-24.4) | 50.0 | 21.4 | 28.6 | 57.1/28.6/14.3 | 84.6/15.4 | 64.3/0.0/35.7 |
| 6 | F (n=10) | 69.5 (59.0-81.0) | 59.3 (52.0-76.0) | 23.7 (16.8-37.1) | 118.9 (110.0-128.5) | 15.9 (13.9-24.9) | 60.0 | 10.0 | 30.0 | 30.0/50.0/20.0 | 60.0/40.0 | 90.0/10.0/0.0 |
|  | M (n=9) | 68.0 (64.0-78.0) | 59.0 (55.0-71.0) | 22.7 (20.8-32.5) | 121.5 (116.0-125.5) | 16.3 (14.5-21.9) | 55.5 | 0.0 | 45.5 | 11.2/44.4/44.4 | 77.8/22.2 | 77.8/11.1/11.1 |
| 7 | F (n=9) | 70.0 (66.0-82.0) | 64.0 (55.0-76.0) | 26.8 (24.5-40.5) | 130.0 (123.5-132.9) | 17.6 (15.0-24.0) | 55.5 | 11.1 | 34.4 | 22.2/44.4/33.3 | 88.8/11.2 | 55.5/33.3/11.2 |
|  | M (n=16) | 70.5 (65.0-85.0) | 59.5 (55.0-76.0) | 26.6 (21.1-44.5) | 123.8 (116.0-140.6) | 17.0 (13.9-24.6) | 56.3 | 12.5 | 31.2 | 33.4/53.3/13.3 | 93.8/6.2 | 68.8/18.8/12.4 |
| 8 | F (n=16) | 74.5 (67.0-91.0) | 60.5 (53.0-80.0) | 32.1 (24.1-48.5) | 132.5 (124.1-142.0) | 18.3 (15.1-26.4) | 50.0 | 18.8 | 31.2 | 6.7/86.6/6.7 | 56.3/43.7 | 93.8/0.0/6.2 |
|  | M (n=16) | 75.0 (62.0-97.0) | 63.0 (53.0-85.0) | 28.4 (19.8-63.1) | 128.2 (118.3-153.5) | 17.6 (11.4-26.8) | 50.0 | 18.8 | 31.2 | 37.4/56.3/6.3 | 73.3/26.7 | 81.3/0.0/18.7 |
| 9 | F (n=12) | 77.0 (68.0-103.0) | 64.0 (53.0-82.0) | 36.9 (27.9-63.2) | 141.6 (134.9-149.8) | 18.4 (14.5-30.0) | 58.3 | 16.7 | 25.0 | 41.7/50.0/8.3 | 100.0/0.0 | 100.0/0.0/0.0 |
|  | M (n=12) | 80.6 (71.0-97.0) | 70.0 (58.0-90.5) | 38.5 (27.9-59.6) | 137.3 (125.7-152.2) | 20.1 (16.1-28.3) | 33.3 | 41.7 | 25.0 | 33.3/66.6/0.0 | 91.7/8.3 | 75.0/16.7/8.3 |
| 10 | F (n=14) | 79.0 (74.0-99.0) | 65.5 (57.0-82.2) | 38.5 (30.4-61.7) | 143.0 (137.8-159.5) | 19.2 (15.6-27.3) | 50.0 | 28.6 | 21.4 | 35.7/50.0/14.3 | 57.1/42.9 | 78.6/14.3/7.1 |
|  | M (n=15) | 87.0 (73.0-98.0) | 71.0 (60.0-90.0) | 46.7 (30.5-66.4) | 146.7 (132.6-152.5) | 20.8 (16.1-30.7) | 26.7 | 33.3 | 40.0 | 40.0/53.3/6.7 | 93.3/6.7 | 86.7/13.3/0.0 |
| 11 | F (n=20) | 86.5 (74.0-106.0) | 65.0 (58.0-83.0) | 46.4 (32.4-74.3) | 153.0 (143.0-173.2) | 19.3 (15.5-28.0) | 65.0 | 25.0 | 10.0 | 10.0/80.0/10.0 | 60.0/40.0 | 95.0/5.0/0.0 |
|  | M (n=17) | 88.0 (69.0-101.0) | 73.0 (58.0-85.0) | 47.0 (29.6-63.2) | 149.0 (138.0-161.0) | 20.8 (15.2-27.4) | 35.3 | 35.3 | 29.4 | 11.7/82.4/5.9 | 76.5/23.5 | 94.1/0.0/5.9 |
| 12 | F (n=21) | 88.0 (79.0-108.0) | 66.0 (60.0-84.0) | 48.8 (36.7-72.6) | 156.5 (146.0-165.2) | 20.4 (16.5-29.5) | 66.7 | 19.0 | 14.3 | 33.3/52.4/14.3 | 71.4/28.6 | 61.9/23.8/14.3 |
|  | M (n=20) | 90.0 (74.0-107.0) | 72.0 (62.0-93.0) | 54.1 (33.9-77.5) | 156.5 (142.9-170.5) | 22.2 (16.1-28.8) | 25.0 | 50.0 | 25.0 | 30.0/60.0/10.0 | 80.0/20.0 | 85.0/10.0/5.0 |
| 13 | F (n=14) | 89.0 (75.0-123.0) | 67.5 (58.0-100.0) | 51.2 (35.0-92.9) | 154.6 (145.3-167.1) | 20.8 (14.6-38.4) | 71.4 | 7.1 | 21.5 | 21.4/78.6/0.0 | 50.0/50.0 | 57.1/35.8/7.1 |
|  | M (n=17) | 91.0 (81.0-128.0) | 71.0 (62.0-100.0) | 59.1 (42.2-112.6) | 165.6 (152.1-180.2) | 20.4 (15.1-38.5) | 58.8 | 5.9 | 35.3 | 29.4/70.6/0.0 | 81.3/18.7 | 82.4/17.6/0.0 |
| 14 | F (n=19) | 90.0 (81.5-106.0) | 69.5 (58.0-90.0) | 53.2 (43.4-73.5) | 160.5 (151.7-171.5) | 19.3 (16.1-29.6) | 68.4 | 26.3 | 5.3 | 36.8/63.2/0.0 | 36.8/63.2 | 78.9/15.8/5.3 |
|  | M (n=11) | 94.0 (71.0-123.0) | 72.0 (67.0-101.0) | 61.7 (46.0-106.9) | 171.5 (162.5-189.5) | 20.9 (17.1-36.1) | 63.6 | 27.3 | 9.1 | 9.1/90.9/0.0 | 81.8/18.2 | 81.8/0.0/18.2 |
| 15 | F (n=15) | 88.0 (82.0-99.0) | 68.0 (59.0-81.0) | 52.5 (43.5-63.4) | 158.5 (156.0-175.5) | 20.3 (16.9-24.5) | 93.3 | 6.7 | 0.0 | 40.0/60.0/0.0 | 42.8/57.2 | 86.8/6.6/6.6 |
|  | M (n=23) | 94.0 (80.0-112.0) | 75.5 (60.0-96.0) | 62.6 (43.5-93.3) | 173.1 (156.0-184.5) | 21.2 (17.9-30.3) | 78.3 | 8.7 | 13.0 | 52.2/39.1/8.7 | 52.2/47.8 | 82.6/8.7/8.7 |
| 16 | F (n=23) | 94.0 (82.0-138.5) | 70.0 (61.0-137.0) | 57.1 (41.0-146.1) | 157.8 (143.5-172.2) | 23.1 (17.5-49.3) | 73.9 | 21.7 | 4.4 | 34.8/52.2/13.0 | 39.1/60.9 | 65.2/21.7/13.1 |
|  | M (n=22) | 97.0 (83.0-121.0) | 76.0 (64.0-100.0) | 71.1 (49.4-110.8) | 176.0 (164.2-186.5) | 22.8 (18.2-35.8) | 63.6 | 27.3 | 9.1 | 22.7/68.2/9.1 | 77.3/22.7 | 77.3/4.5/18.2 |
| 17 | F (n=19) | 91.5 (84.0-110.0) | 72.0 (59.0-87.0) | 60.6 (50.7-76.1) | 163.0 (149.5-170.0) | 23.0 (17.9-28.0) | 73.7 | 26.3 | 0.0 | 0.0/84.2/15.8 | 21.1/78.9 | 84.2/10.5/5.3 |
|  | M (n=15) | 92.0 (85.0-111.0) | 73.5 (64.0-91.0) | 66.5 (50.5-96.0) | 176.2 (160.5-186.8) | 22.2 (17.8-30.0) | 80.0 | 13.3 | 6.7 | 46.7/40.0/13.3 | 73.3/26.7 | 66.7/26.7/6.6 |

The hip-to-height ratio was the best anthropometric index associated with BMI (r = 0.78, *p* = 1.3×10^−86^) vs the waist-to-hip ratio (r = 0.13, *p* = 0.007) and the waist-to-height ratio (r = 0.32, *p* = 0.001) (Supplementary Fig. 1). As expected in growing children, BMI values increased on average from 5 to 17 years (Fig. 1A). The percentage of overweight-obesity decreased from 5 to 17 years (from 50% to 25%, respectively) with the exemption of boys from 9 to 12 years old (*n* = 64) that reached the highest prevalence of overweight-obesity (∼70%) (Fig. 1B).

**Figure 1.**
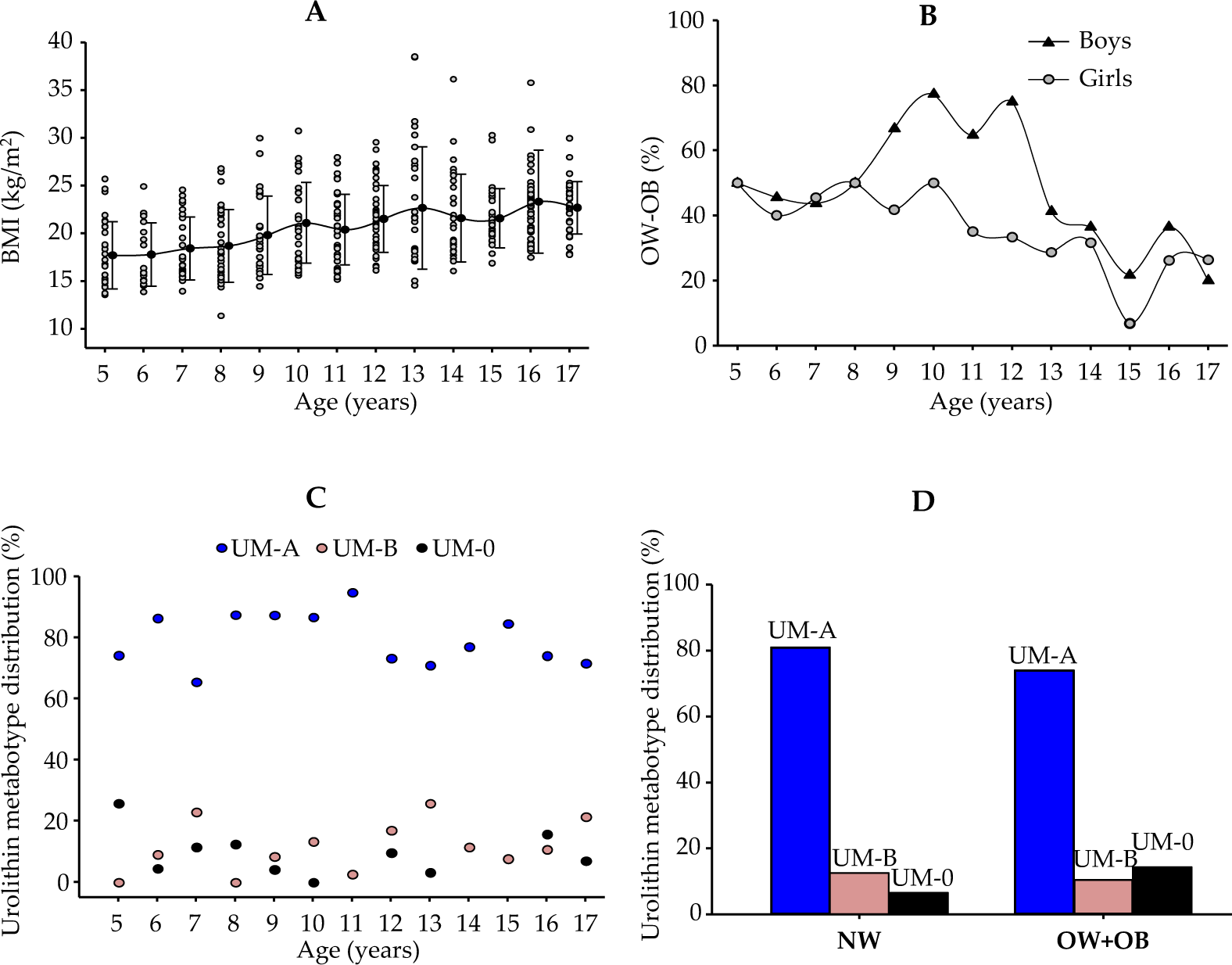
Distribution (%) of (**A**) BMI, (**B**) overweight–obesity (OW–OB) and (**C**) urolithin metabotypes in the cohort (*n* = 415) from 5 to 17 years. (**D**) Distribution of urolithin metabotypes (UM-A, UM-B and UM-0) in normoweight (NW) and OW+OB children.

Regarding the SNPs analysed, after correcting for multiple tests, three SNPs did not satisfy the Hardy-Weinberg equilibrium (HWE) (rs1801253, rs5082, rs11868035) (Supplementary Table 1). The rest of the SNPs were in equilibrium and were close to European frequencies. In the case of these three SNPs, we can speculate that the reason could be the association established by the presence of certain consanguinity (several sibling groups in the cohort). Nevertheless, this did not affect our results, since we aimed to compare variables from different domains and not only in estimating the particular effect of a single SNP. Overall, it is not absolutely necessary to have HWE in our approach, i.e., to estimate odds ratios of an MCA dimension and rank the predictors.

The distribution of urolithin metabotypes in this cohort (Fig. 1C) revealed that both a lower prevalence of UM-A and a higher occurrence of UM-0 were associated with an increased percentage of overweight-obesity after bivariate analysis (*p* = 0.015) (Fig. 1D).

No significant association between physical activity and overweight-obesity distribution was found (results not shown). On the contrary, there were many overweight-obese children with high physical activity, which should be explained as a consequence of their overweight-obesity status (results not shown). Regarding the diet, the KIDMED scores showed mean values of 6.9 ± 2.1 for the entire cohort (Supplementary Table 2) and ranged from the lowest value of 5.7 ± 1.4 in 15-year-old girls to the highest value of 8.8 ± 2.2 in 6-year-old boys (Fig. 2A). However, no significant differences were found between boys and girls as well as through the range of age (Fig. 2A). Besides, no significant association was found between overweight-obesity and KIDMED in this group (results not shown).

**Figure 2.**
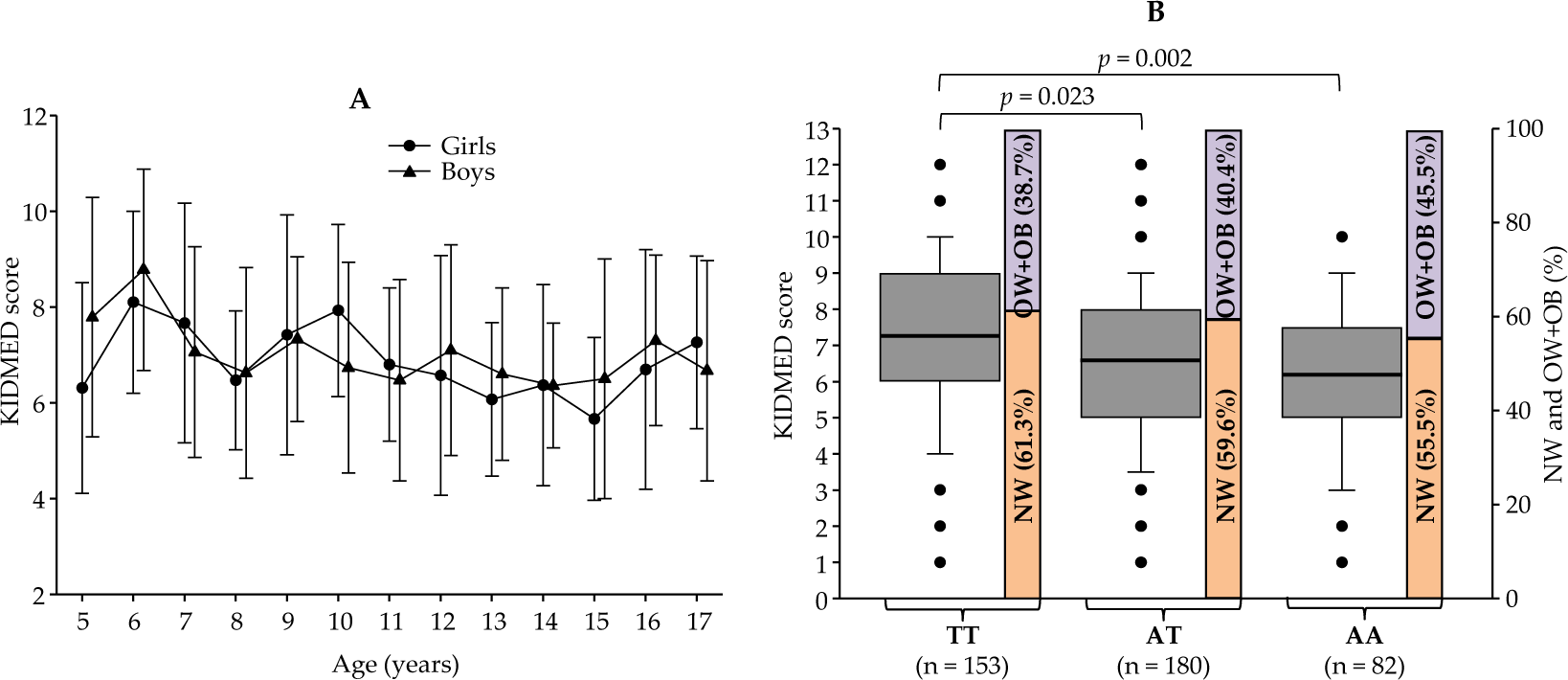
(A) KIDMED scores in girls (●) and boys (▴) from 5 to 17 years. Results are expressed as mean ± SD. (B) KIDMED scores (box plots) as a function of the genotypes (TT, AT and AA) of the rs9939609 SNP in the FTO gene. Significant differences are shown after the Kruskal Wallis test. Bar charts show the proportion of normoweight (NW) and overweight-obesity (OW+OB) in the cohort depending on the rs9939609-*FTO* genotype of the individuals.

We next explored the potential association of both adherence to the Mediterranean diet and percentage of overweight-obesity with the well-known obesity risk allele A of the rs9939609 SNP in the *FTO* gene. Figure 2B shows the KIDMED scores and the percentage of normoweight and overweight-obesity in children depending on their genotype TT, AT or AA. Remarkably, the adherence to the Mediterranean diet was significantly lower in children with the risk-associated genotypes AT and AA vs the TT genotype (Fig. 2B). Although the proportion of overweight-obesity was the highest for the AA genotype (45.5%), however, the difference vs that of the TT genotype (38.7%) did not reach statistical significance (Fig. 2B). Once again, many exemptions prevented the usefulness of the combination of KIDMED scores and rs9939609 genotypes as unique predictors of overweight-obesity in this cohort.

Therefore, although both the urolithin metabotypes and rs9939609 SNP-*FTO* could partially contribute to the overweight-obesity distribution as independent predictor variables in this cohort of children and adolescents; however, all the possible SNP-SNP interactions together with the rest of variables had not been taken into account. Therefore, we next developed an ordinal logistic model to identify the consortium of variables that could estimate the odds ratios of the overweight-obesity distribution in this cohort.

### An Ordinal Logistic Model to Identify the Consortium of Variables Associated with Overweight-Obesity

In this model, we used as the ordinal response the normoweight, overweight and obesity classification for children, based on sex and age to estimate the odds ratios (and the corresponding 95% confidence intervals) for different predictor variables, as well as their relative importance in the prediction of the response. In this holistic approach, we used as predictors the ‘Ethnicity’, ‘Urolithin metabotypes’, ‘KIDMED score’, ‘Physical activity’, and genetic polymorphisms (44 SNPs were finally included and compressed into 15 MCA dimensions, hereafter termed ‘SNP.Dim.’), together with sex and age. Although the distribution of normoweight, overweight and obesity WHO-based categories was apparently adjusted by sex and age, we still observed a trend for decreasing average age when moving from normoweight, overweight and obese children, as well as enrichment in boys in the same order. Therefore, we also included these variables in the model.

Figure 3 displays the predictors used in the ordinal logistic model ranked by their apparent importance, as measured by their χ^2^-df score. The model was highly significant (*p* < 0.0001), yielding the components ‘Age’, and ‘SNP.Dim.14’ as the two most important predictors. Therefore, in the prediction of overweight-obesity in this population, apart from ‘Age’ and ‘Sex’, the most critical contributing variables were SNPs (through the variable SNP.Dim.14), followed by ‘Ethnicity’, ‘Urolithin metabotype’ and ‘KIDMED’. The ‘Physical activity’ seemed to be irrelevant in this sample.

**Figure 3.**
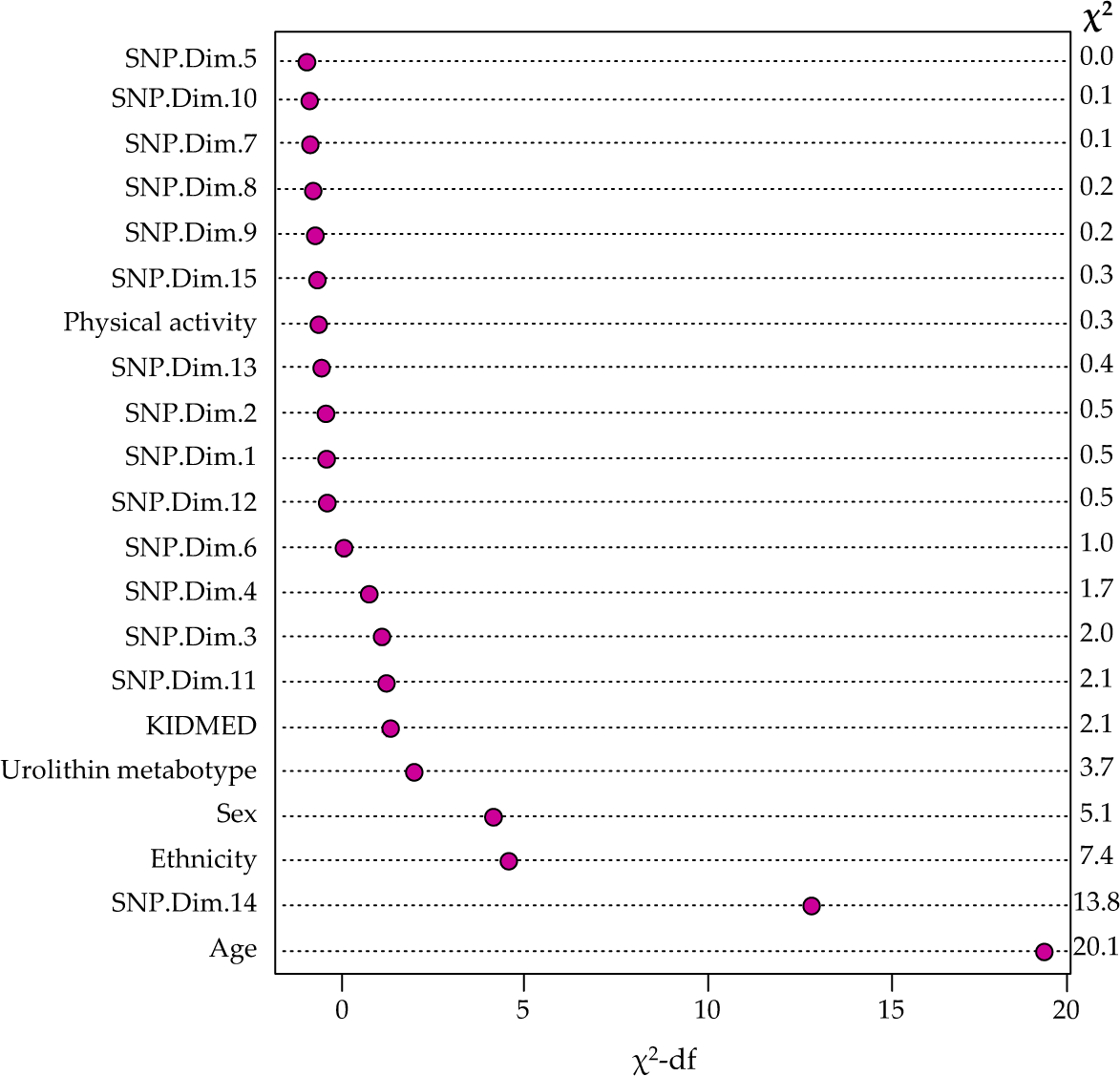
Apparent predictor importance in the ordinal logistic model. Variables are ranked by relevance (based on their χ^2^-df score). The χ^2^ of the Wald test for each predictor is also shown

We next applied a fast-backwards approach based on AIC to obtain a reduced model and estimate the corresponding βs and ORs (as well as their 95% CI). The variables kept were ‘Sex’, ‘Age’, ‘Urolithin metabotype’, ‘KIDMED’, and the genetic components ‘SNP.Dim.3’, ‘SNP.Dim.11’, and ‘SNP.Dim.14’ (Table 2). The variable ‘Ethnicity’ was not retained in the simplified model, probably due to its high complexity (4 levels). Our results reveal that being a boy either with UM-B or UM-0 and having a higher SNP.Dimension.14, all increased the chances of overweight-obesity in this study population. On the contrary, ageing, better adherence to the Mediterranean diet (higher KIDMED score), and being UM-A was associated with lower probabilities of being overweighed-obese. Remarkably, all these variables operated additively to build the final probability of overweight-obesity for each subject.

**Table 2.** Beta estimates (βs), odds ratios (ORs), and their corresponding confident intervals (CIs). *SNP.Dim, dimensions (consortium) of SNPs obtained after Multiple Component Analysis; UM-B and UM-0, urolithin metabotype B and 0, respectively.

| Variables* | βs (CIs) | ORs (CIs) |
| --- | --- | --- |
| Sex (boy) | 0.51 (0.10, 0.918) | 1.67 (1.11, 2.5) |
| Age | -0.12 (-0.17, -0.0664) | 0.88 (0.83, 0.93) |
| UM-B | 0.21 (-0.40, 0.837) | 1.24 (0.66, 2.31) |
| UM-O | 0.68 (0.02, 1.34) | 1.98 (1.03, 3.81) |
| KIDMED | -0.06 (-0.16, 0.0256) | 0.93 (0.85, 1.03) |
| SNP.Dim.3 | 1.05 (0.03, 2.07) | 2.86 (1.03, 7.94) |
| SNP.Dim.11 | 1.18 (-0.01, 2.37) | 3.25 (0.98, 10.7) |
| SNP.Dim.14 | 2.32 (1.08, 3.55) | 10.1 (2.94, 35) |

Figure 4 shows the R^2^ values of the 24 SNPs that significantly contributed to SNP.Dim.14 (*p* < 0.05). The most contributing SNP to this consortium was rs1801253-*ADRB1*, as well as other obesity risk-associated SNPs (*UCP2, ADIPOQ, LEP, MC4R*, etc.). However, there were other SNPs with less known involvement in obesity. Table 3 shows all the SNPs contributing to SNP.Dim.14 with their definitions and main processes in which they are involved.

**Table 3.**
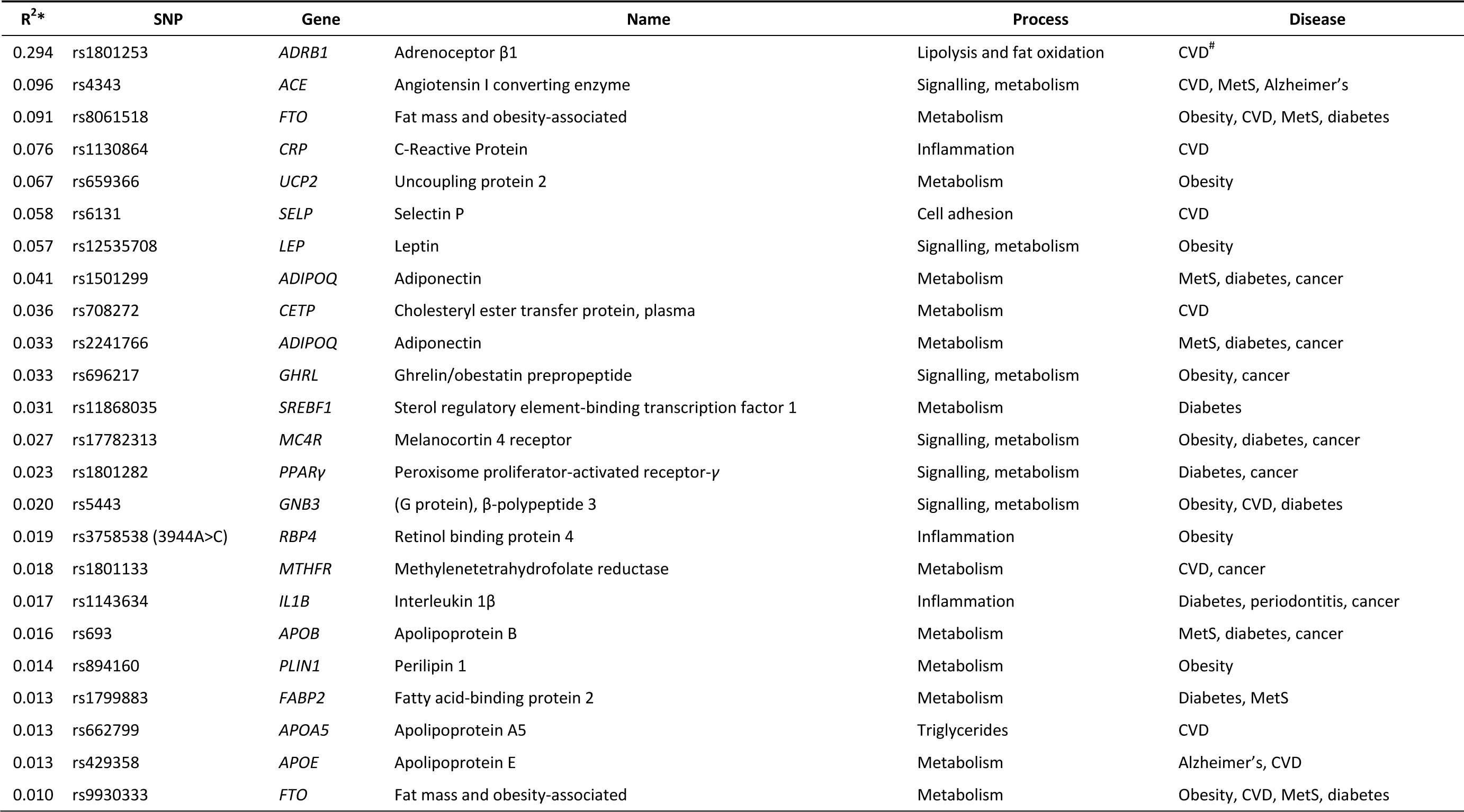
The consortium of single-nucleotide polymorphisms (SNPs) integrated into the MCA dimension SNP.Dim.14. *R^2^, Contribution of the SNPs to Dim.14 according to Figure 3. ^#^CVD, cardiovascular disease; MetS, metabolic syndrome

| R<sup>2</sup>* | SNP | Gene | Name | Process | Disease |
| --- | --- | --- | --- | --- | --- |
| 0.294 | rs1801253 | *ADRB1* | Adrenoceptor β1 | Lipolysis and fat oxidation | CVD<sup>#</sup> |
| 0.096 | rs4343 | *ACE* | Angiotensin I converting enzyme | Signalling, metabolism | CVD, MetS, Alzheimer's |
| 0.091 | rs8061518 | *FTO* | Fat mass and obesity-associated | Metabolism | Obesity, CVD, MetS, diabetes |
| 0.076 | rs1130864 | *CRP* | C-Reactive Protein | Inflammation | CVD |
| 0.067 | rs659366 | *UCP2* | Uncoupling protein 2 | Metabolism | Obesity |
| 0.058 | rs6131 | *SELP* | Selectin P | Cell adhesion | CVD |
| 0.057 | rs12535708 | *LEP* | Leptin | Signalling, metabolism | Obesity |
| 0.041 | rs1501299 | *ADIPOQ* | Adiponectin | Metabolism | MetS, diabetes, cancer |
| 0.036 | rs708272 | *CETP* | Cholesteryl ester transfer protein, plasma | Metabolism | CVD |
| 0.033 | rs2241766 | *ADIPOQ* | Adiponectin | Metabolism | MetS, diabetes, cancer |
| 0.033 | rs696217 | *GHRL* | Ghrelin/obestatin prepropeptide | Signalling, metabolism | Obesity, cancer |
| 0.031 | rs11868035 | *SREBF1* | Sterol regulatory element-binding transcription factor 1 | Metabolism | Diabetes |
| 0.027 | rs17782313 | *MC4R* | Melanocortin 4 receptor | Signalling, metabolism | Obesity, diabetes, cancer |
| 0.023 | rs1801282 | *PPARγ* | Peroxisome proliferator-activated receptor-γ | Signalling, metabolism | Diabetes, cancer |
| 0.020 | rs5443 | *GNB3* | (G protein), β-polypeptide 3 | Signalling, metabolism | Obesity, CVD, diabetes |
| 0.019 | rs3758538 (3944A>C) | *RBP4* | Retinol binding protein 4 | Inflammation | Obesity |
| 0.018 | rs1801133 | *MTHFR* | Methylenetetrahydrofolate reductase | Metabolism | CVD, cancer |
| 0.017 | rs1143634 | *IL1B* | Interleukin 1β | Inflammation | Diabetes, periodontitis, cancer |
| 0.016 | rs693 | *APOB* | Apolipoprotein B | Metabolism | MetS, diabetes, cancer |
| 0.014 | rs894160 | *PLIN1* | Perilipin 1 | Metabolism | Obesity |
| 0.013 | rs1799883 | *FABP2* | Fatty acid-binding protein 2 | Metabolism | Diabetes, MetS |
| 0.013 | rs662799 | *APOA5* | Apolipoprotein A5 | Triglycerides | CVD |
| 0.013 | rs429358 | *APOE* | Apolipoprotein E | Metabolism | Alzheimer's, CVD |
| 0.010 | rs9930333 | *FTO* | Fat mass and obesity-associated | Metabolism | Obesity, CVD, MetS, diabetes |

**Figure 4.**
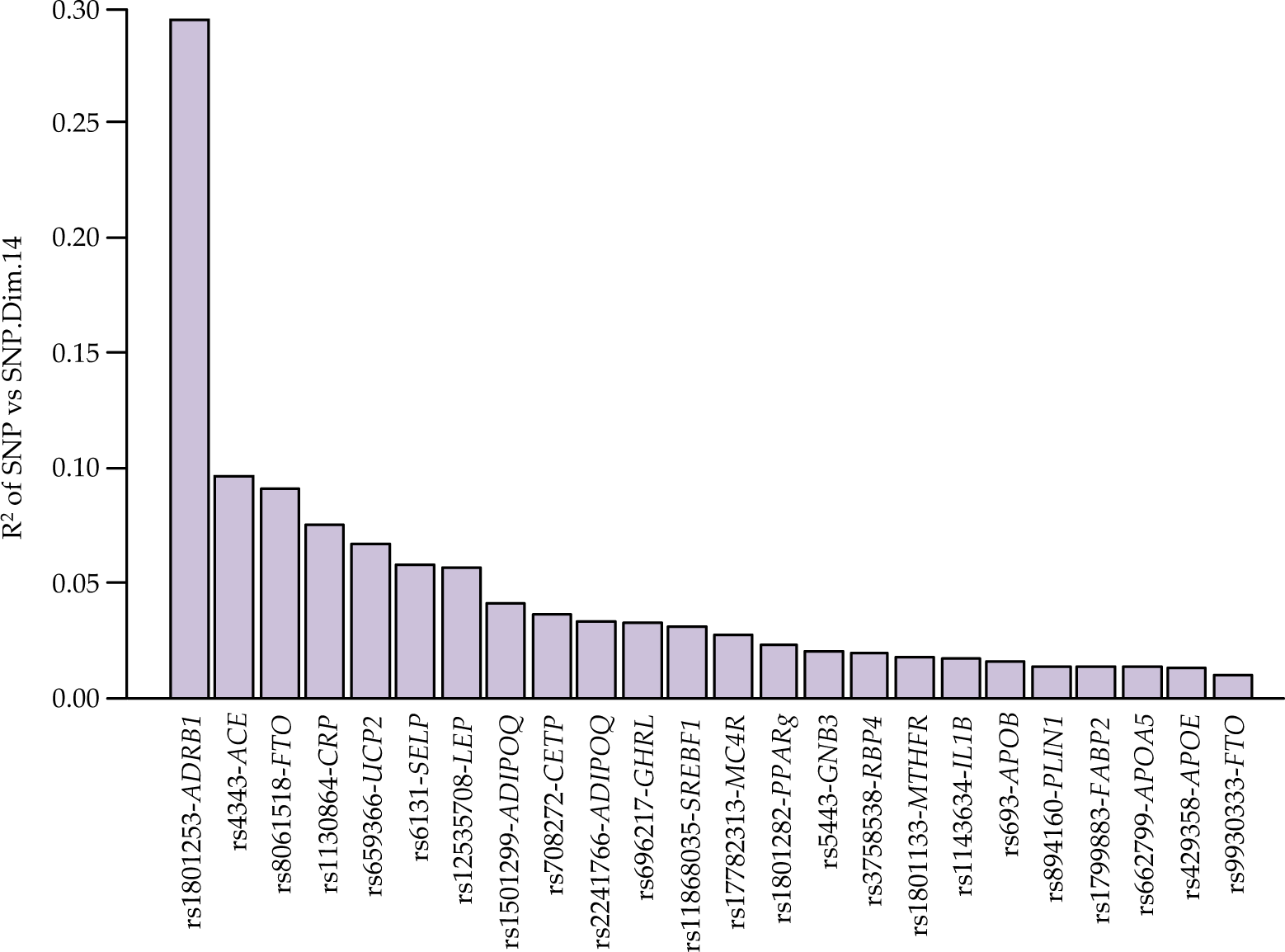
Description of components of SNP Dimension-14. Significant R^2^ (*p* < 0.05) for each SNP within the SNP Dimension-14 are shown.

### Model Validation

We used bootstrap to validate the model (Table 4). The model showed some degree of overfitting, reflected in a decrease of the indexes after correction for optimism. The discriminative capacity of the model was modest, although not negligible according to the Dxy, R^2^ indexes (it must be taken into account that the typical values of R^2^ in ordinal models are much lower than those observed in linear regression models), and the Brier score of 0.23. The calibration showed some degree of shrinkage, as seen from the deviation of the intercept and slope from 0 and 1, respectively. E_max_, the maximum calibration error in predicting *p* (Y > normal), was 0.11, showing some degree of miscalibration. Overall, we obtained a significant model that still showed predictive power in external data.

**Table 4.**
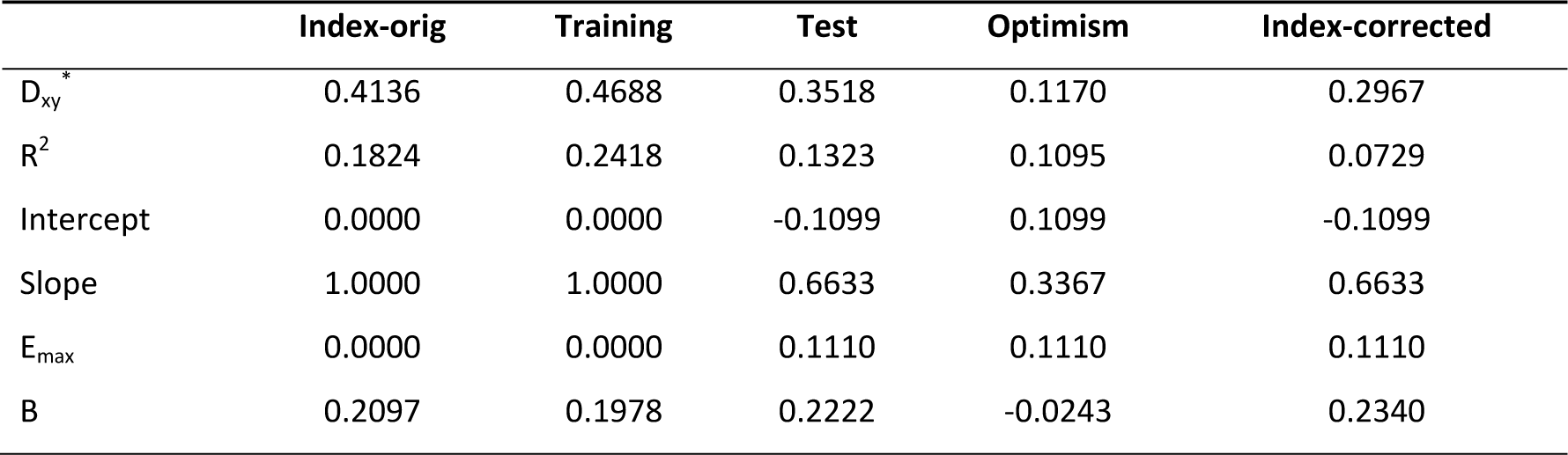
Validation of the model using Bootstrap. **\***D_xy_ = bias-corrected Somers rank correlation coefficient that goes from −1 to 1; R^2^ = Nagelkerke R^2^ that goes from 0 to 1; Intercept and Slope of a logistic calibration equation (should be 0 and 1, respectively, for a perfect fit); E_max_ = maximum calibration error for *p* (Y = 0) based on the linear-logistic recalibration; B = Brier’s quadratic probability score, which goes from 0 (the best score) to 1 (the worst score).

|  | Index-orig | Training | Test | Optimism | Index-corrected |
| --- | --- | --- | --- | --- | --- |
| $D_{xy}^*$ | 0.4136 | 0.4688 | 0.3518 | 0.1170 | 0.2967 |
| $R^2$ | 0.1824 | 0.2418 | 0.1323 | 0.1095 | 0.0729 |
| Intercept | 0.0000 | 0.0000 | -0.1099 | 0.1099 | -0.1099 |
| Slope | 1.0000 | 1.0000 | 0.6633 | 0.3367 | 0.6633 |
| $E_{max}$ | 0.0000 | 0.0000 | 0.1110 | 0.1110 | 0.1110 |
| B | 0.2097 | 0.1978 | 0.2222 | -0.0243 | 0.2340 |

## Discussion

The present study shows that the probabilities of childhood overweight-obesity cannot be explained by specific isolated variables (that is, one or a few SNPs, or just physical activity, or diet, etc.), but by complex, multifactorial associations of environmental and genetic components., Besides, a genetic score, widely used in the literature^36^, and understood as the theoretical equal-additive contribution of every single SNP could not explain the distribution of overweight-obesity in our cohort. In this regard, our analysis adds significance to previous studies that have associated the genetic background of adult individuals with the adherence to the Mediterranean diet and some markers related to obesity and metabolic syndrome^36,37,38^. However, to the best of our knowledge, there are no previous studies that explore the occurrence of overweight-obesity in children and adolescents by estimating the odds ratios for the predictor variables ‘urolithin metabotypes’ as gut microbiota biomarkers, ‘age’, ‘sex’, ‘adherence to the Mediterranean diet’, and an identified consortium of 24 SNPs from 22 genes, mainly related to obesity and cardiometabolic diseases.

The purpose of the ordinal logistic regression model was to estimate the odds ratios of the different explanatory variables and rank them by importance, rather than a tool to predict the probability of overweight and obesity of new individuals. There are in the literature several previous reports that model the probabilities of overweight and obesity in children through ordinal regression models^39,40,41,42,43^. However, to the best of our knowledge, this is the first time that this type of model is used with such a wide set of predictors comprising different putatively influencing domains (genetic, diet, exercise, microbiome urolithin metabotype, and ethnicity, in addition to sex and age) and ranks them according to their relative importance. The χ^2^-df score returned the following importance ranking: age > SNP.Dim.14 > ethnicity > sex > urolithin metabotype > KIDMED; while physical activity seemed to have a negligible importance. The use of an AIC-based fast-backwards reduction of the model removed ‘Ethnicity’ from the final model, probably because of the large number of categories in this predictor^39^, that would be highly penalised by the AIC criterion. Besides, our sample was mainly of Caucasian-European origin (93.5%), and it is expected that in a sample with a more balanced distribution of ethnicity, this predictor would have high importance. Furthermore, the modest predictive capability of the model suggested the need for additional predictors, which is not unexpected, given the multifaceted aetiology of obesity. Still, the model, derived for estimating purposes, allowed us to rank predictors of different domains by their importance, as well as to estimate odds ratios for them.

The classification of overweight and obesity in adults is rather simple and independent of age and sex (i.e. 25 ≤ BMI < 30 for overweight, and BMI ≥ 30 for obesity). However, the WHO-based definition for overweight-obesity in children and teens uses age- and sex-based percentiles^5^, and a higher BMI does not necessarily correspond with overweight-obesity in growing children. Therefore, using overweight and obesity as response variables instead of BMI values is especially useful in children and adolescents. This is due to the difficulty in comparing them at different ages and of the two sexes, given the remarkable, sex-dependent change in weight and height, during this period of development (i.e., while our model predicted that overweight-obesity decreases upon ageing, the use of BMI yielded the opposite result, masking the real result). Still, sex and age were essential predictors in our model, with estimated ORs of 1.67 for boys, and 0.88 for a one-year increase, respectively, which means an increased percentage of overweighed-obese boys, and a reduction of overweight-obesity upon ageing.

The gut microbiota contributes to the pathophysiology of obesity^3^, and a recent report shows the role of gut microbial metabolites in the expression of the *microRNA-181* family, which regulates white adipose tissue inflammation and obesity in children^14^. However, the gut microbiota, as a predictor of childhood obesity, has been scarcely approached and not usually considered together with SNPs and other variables. In a targeted approach, among a few analysed microbial groups, Scheepers et al. reported that the *Bacteroides fragilis* group was associated with childhood weight development^44^. Later, Rampelli et al. connected the onset of obesity in children to an increase of Proteobacteria and a decrease of Clostridiaceae and Ruminococcaceae^45^. In the same line, Bai et al. described that the Proteobacteria phylum had significantly enriched OTUs for higher BMI levels in a cohort of children from the American Gut Project^46^. This is relevant since the microbiota-associated with UM-B is enriched in Proteobacteria^24^. Interestingly, Nirmalkar et al. reported that the Coriobacteriaceae family was 3-fold more abundant in obese children and adolescents than normoweight^47^. Specifically, the genus *Collinsella* was more abundant in obese adolescents, a microbial group that could be related to the endothelial dysfunction^47,48^. Overall, all these results are remarkable since we recently reported that UM-B was enriched in the Coriobacteriaceae family, which was positively correlated with blood total-cholesterol, LDL-cholesterol, and BMI in adults^24^. Although we had previously observed a trend between UM-B occurrence and overweight-obesity in adult individuals, however, we could not establish a definitive link probably due to the lack of other interacting factors such as those included in the present study (SNPs, diet, etc.)^25,49^. Therefore, the Coriobacteriaceae family, and probably the Proteobacteria phylum, more abundant in obese children as well as in UM-B, could be the link between UM-B occurrence and overweight-obesity, which suggests that the microbiota associated with urolithin metabotypes could contribute in the prediction of the probability of being overweighed or obese. Regarding UM-0, its occurrence in the population is approximately constant (∼10%), although there is both a higher occurrence and variability in the childhood^25^. The microbiota associated with UM-0 has been reported to show lower diversity than UM-B and UM-A, which could be indicative of an obesity-prone microbiota^4^. Nevertheless, we cannot exclude a possible shift of metabotype in children from UM-0 to either UM-A or UM-B determined by ageing^25^ or after ellagitannin-rich diets as previously described in adults^20^.

Regarding the possible health implications of belonging to one or another metabotype, we have recently reported that urolithin metabotypes determined the different restoration capacity of the gut microbiota and the anthropometric values (weight, waist and hip) of healthy women up to 12 months after delivery^50^. The gut microbiota of pregnant women is in dysbiosis, which persists at least 1 month after delivery. We observed that the gut microbiota associated with UM-B was more resilient than that of UM-A, which would have negative implications in the dysbiotic-prone UM-B. In contrast, the gut microbiota of UM-A women progressively became normal during the year after childbirth. Therefore, we suggested that the determination of urolithin metabotypes in pregnant and lactating women could be a useful tool to predict their predisposition to the recovery of the gut microbiota and anthropometric values, significantly altered during pregnancy and after childbirth^50^.

Many studies describe the association, or lack of association, of specific SNPs with obesity. The rs9939609 SNP-*FTO* has been reported to confer a predisposition to obesity by regulating the control of food intake and food choice, suggesting a link to a hyperphagic phenotype or a preference for energy-dense foods in Scottish children^51^. This agrees with the connection between the low adherence to the Mediterranean diet and the risk allele A of rs9939609 SNP-*FTO* in our cohort. However, we did not observe a clear association of this SNP with overweight-obesity, but only a trend with many exemptions, in agreement with other studies where some SNPs, previously reported to be involved in obesity, such as the rs17782313-*MC4R* and rs9939609-*FTO*, exerted weak effects and very scarce contribution to obesity in 773 pre-pubertal Portuguese children^52^. In the present study, a consortium of 24 SNPs was identified as the second contributing predictor to overweight-obesity in our cohort. The rs1801253 SNP (also called Arg389Gly), located in the gen *ADRB1*, was the most contributing SNP within this consortium. It is known the involvement of the *ADRB1* gene polymorphisms in cardiovascular diseases^53^ but also in obesity^54,55^.

The *ADRB1* gene codifies a G-coupled protein (the β1-adrenergic receptor) that binds the catecholamines epinephrine and norepinephrine and controls sympathetic responses in the heart, kidney and adipocytes. Interestingly, Dionne et al. reported that the rs1801253 SNP-*ADRB1* was associated with higher body weight and BMI in a cohort of Caucasian women (*n* = 931)^54^. In the same line, Aradillas-García et al. reported that the rs1801253 SNP-*ADRB1*, but not the Trp64Arg *ADRB3*, was associated with obesity in Mexican children^56^. The connection between *ADBR1* SNPs and obesity could rely on the catecholamines, which are considered significant lipolysis regulators^57^ and affect differentiation and proliferation of adipocytes^58^. In this regard, Lee et al.^59^ established the association between impaired urinary epinephrine and norepinephrine excretion and obesity, insulin resistance, and metabolic syndrome in a cohort of 577 Chinese subjects.

Overall, all the above highlights again the need to consider SNPs consortia, interacting with other variables, instead of few SNPs in those studies aimed to associate SNPs with obesity. We are aware that the present study is an exploratory validation for a proof-of-concept, i.e., an ordinal logistic model that associates child overweight-obesity with a consortium of SNPs potentially interacting with the urolithin metabotypes-associated microbiota, adherence to the Mediterranean diet, age, and sex. Although we claim for the rationale of our approach and its potential usefulness, however, our results should be confirmed with additional research. We also acknowledge some limitations that should be considered in further studies, which also could improve its prediction capability. For example, it would be interesting to include other possible variables, such as the detailed composition and functionality of the individuals’ gut microbiomes, and dietary interventions to evaluate not only associations but also individuals’ responses. The latter would be even better than the use of validated questionnaires. Besides, a higher number of SNPs (or many SNPs associated with a specific gene) should be explored, especially in children from other geographical origins and ethnicities. Also, the inclusion of serobiochemical variables and traits related to obesity and its comorbidities (blood lipid profile, blood pressure, glucose homeostasis, etc.) could yield relevant information. Finally, a validation cohort (i.e., a parallel-group with all the children either normoweight or obese) should also be considered in further studies to confirm our model fully.

## Conclusions

The present research highlights the need for a holistic approach to unravel the predictors of overweight-obesity in children. Our results confirm, in agreement with the multifaceted aetiology of obesity, the link of childhood overweight-obesity to multifactorial associations of environmental and genetic components. The ordinal logistic model revealed that child overweight-obesity prevalence was related to being a young boy with either UM-B or UM-0, low KIDMED score and high contribution of a consortium of 24 SNPs, being rs1801253-*ADRB1*, rs4343-*ACE*, rs8061518-*FTO*, rs1130864-*CRP*, rs659366-*UCP2*, rs6131-*SELP*, rs12535708-*LEP*, rs1501299-*ADIPOQ*, rs708272-*CETP* and rs2241766-*ADIPOQ* the top-ten contributing SNPs. Therefore, it is of particular relevance the evaluation of interactive SNPs consortia along with the stratification of the children according to their urolithin metabotypes, which could be early biomarkers, in the case of UM-B and UM-0, of a dysbiotic-prone obesity-associated microbiota.

## Data Availability

There are no external datasets or supplementary material online at other repositories that pertain to this manuscript.

## Data availability

All data generated or analysed during this study are included in this published article (and its Supplementary Information files).

## Acknowledgments

This research was funded by the Project AGL2015-64124-R (MINECO, Spain). A.C.-M. is the holder of a FPI predoctoral grant from MINECO (Spain). The authors thank Ana Ramírez, Susana Molina and M. Isabel Espinosa from the GENYAL platform (IMDEA-Food, Madrid, Spain) for their help in the genotyping of the students. The authors are also grateful to Carlos Ortiz from the public primary school ‘CEIP Jara Carrillo’ (Alcantarilla, Murcia, Spain), as well as to Mateo Moya, M. Teresa Saavedra and Ginés Bueno from the public high school ‘IES Alcántara’ (Alcantarilla, Murcia, Spain) for being so well disposed towards this project and facilitating the recruitment process. We especially appreciate the participation of the students in this research and the willingness of their parents to make this possible. Finally, we also thank our colleagues R. García-Villalba, M. Romo-Vaquero and M.A. Núñez-Sánchez for their collaboration in the recruitment process.

## Author Contributions

A.C.-M and G.C. contributed equally to this work. Conceptualization, J.C.E; Methodology, Software and Validation, J.C.E, G.C. and A.C.-M.; Funding Acquisition, J.C.E., M.V.S.; Original drafting, J.C.E, Writing–Review & Editing, G.C., A.C.-M. M.V.S., J.C.E.

## References

1. Head, G.A. Cardiovascular and metabolic consequences of obesity. Front. Physiol. 6, 32 (2015).

2. Meldrum, D.R., Morris, M.A. & Gambone, J.C. Obesity pandemic: causes, consequences, and solutions-but do we have the will? Fertil. Steril. 107, 833–839 (2017).

3. Cani, P.D. Human gut microbiome: hopes, threats and promises. Gut. 67, 1716–1725 (2018).

4. Le Roy, C.I. et al. Dissecting the role of the gut microbiota and diet on visceral fat mass accumulation. Sci. Rep. 9, 9758 (2019).

5. World Health Organization (WHO). Growth reference 5-19 years. https://www.who.int/growthref/who2007_bmi_for_age/en/

6. Lopez, R.B., Heatherton, T.F. & Wagner, D.D. Media multitasking is associated with higher risk for obesity and increased responsiveness to rewarding food stimuli. Brain Imaging Behav. (2019).

7. Frayling, T.M. et al. A common variant in the FTO gene is associated with body mass index and predisposes to childhood and adult obesity. Science. 316, 889–894 (2007).

8. Fawcett, K.A. & Barroso, I. The genetics of obesity: FTO leads the way. Trends Genet. 26, 266–274 (2010).

9. Hess, M.E. & Brüning, J.C. The fat mass and obesity-associated (FTO) gene: obesity and beyond? Biochim. Biophys. Acta. 1842, 2039–2047 (2014).

10. Frayling, T.M. & Ong, K. Piecing together the FTO jigsaw. Genome Biol. 12, 2011–2012 (2011).

11. Sachidanandam, R. et al. A map of human genome sequence variation containing 1.42 million single nucleotide polymorphisms. Nature. 409, 928–933 (2001).

12. Vijay-Kumar, M. et al. Metabolic syndrome and altered gut microbiota in mice lacking Toll-like receptor 5. Science. 328, 228–231, (2010).

13. Biesiekierski, J.R., Jalanka, J. & Staudacher, H.M. Can gut microbiota composition predict response to dietary treatments? Nutrients. 11, 1134 (2019).

14. Virtue, A.T. et al. The gut microbiota regulates white adipose tissue inflammation and obesity via a family of microRNAs. Sci. Transl. Med. 11, eaav1892 (2019).

15. Miller, L.M. et al. Being overweight or obese is associated with harboring a gut microbial community not capable of metabolizing the soy isoflavone daidzein to O-desmethylangolensin in peri- and post-menopausal women. Maturitas. 99, 37–42 (2017).

16. Espín, J.C.; González-Sarrías, A. & Tomás-Barberán, F.A. The gut microbiota: a key factor in the therapeutic effects of (poly)phenols. Biochem Pharmacol. 139, 82–93 (2017).

17. Tomás-Barberán, F.A. et al. Urolithins, the rescue of “old” metabolites to understand a “new” concept: metabotypes as a nexus among phenolic metabolism, microbiota dysbiosis, and host health status. Mol. Nutr. Food Res. 61, 1500901 (2017).

18. Selma, M.V. et al. The gut microbiota metabolism of pomegranate or walnut ellagitannins yields two urolithin-metabotypes that correlate with cardiometabolic risk biomarkers: comparison between normoweight, overweight-obesity and metabolic syndrome. Clin. Nutr. 37, 897–905 (2018).

19. Frankenfeld, C.L., Atkinson, C., Wähälä, K. & Lampe, J.W. Obesity prevalence in relation to gut microbial environments capable of producing equol or O-desmethylangolensin from the isoflavone daidzein. Eur. J. Clin. Nutr. 68, 526–530 (2014).

20. González-Sarrías, A. et al. Clustering according to urolithin metabotype explains the interindividual variability in the improvement of cardiovascular risk biomarkers in overweight-obese individuals consuming pomegranate: a randomized clinical trial. Mol. Nutr. Food Res. 61, 1600830 (2017).

21. Hazim, S. et al. Acute benefits of the microbial-derived isoflavone metabolite equol on arterial stiffness in men prospectively recruited according to equol producer phenotype: a double-blind randomized controlled trial. Am. J. Clin. Nutr. 103, 694–702 (2016).

22. Selma, M.V. et al. The human gut microbial ecology associated with overweight and obesity determines ellagic acid metabolism. Food Funct. 7, 1769–1774 (2016).

23. Tomás-Barberán, F.A., Selma, M.V. & Espín, J.C. Polyphenols’ gut microbiota metabolites: bioactives or biomarkers? J. Agric. Food Chem. 66, 3593–3594 (2018).

24. Romo-Vaquero, M. et al. Deciphering the human gut microbiome of urolithin metabotypes: association with enterotypes and potential cardiometabolic health implications. Mol. Nutr. Food Res. 63, e1800958 (2019).

25. Cortés-Martín, A. et al. The gut microbiota urolithin metabotypes revisited: the human metabolism of ellagic acid is mainly determined by aging. Food Funct. 9, 4100–4106 (2018).

26. Nuñez-Sánchez, M.A. et al. Targeted metabolic profiling of pomegranate polyphenols and urolithins in plasma, urine and colon tissues from colorectal cancer patients. Mol. Nutr. Food Res. 58, 1199–1211 (2014).

27. Sylvia, L.G., Bernstein, E.E., Hubbard, J.L., Keating, L. & Anderson, E.J. Practical guide to measuring physical activity. J. Acad. Nutr. Diet. 114, 199–208 (2014).

28. Serra-Majem, L. et al. Food, youth and the Mediterranean diet in Spain. Development of KIDMED, Mediterranean Diet Quality Index in children and adolescents. Public Health Nutr. 7, 931–935 (2004).

29. Sanghera, D.K., Bejar, C., Sharma, S., Gupta, R. & Blackett, P.R. Obesity genetics and cardiometabolic health: potential for risk prediction. Diabetes Obes. Metab. 21, 1088–1100 (2019).

30. Fall, T. & Ingelsson, E. Genome-wide association studies of obesity and metabolic syndrome. Mol. Cell. Endocrinol. 382, 740–757 (2014).

31. Yu, Z. et al. Genetic polymorphisms in adipokine genes and the risk of obesity: a systematic review and meta-analysis. Obesity (Silver Spring). 20, 396–406 (2012).

32. Marcos-Pasero, H. et al. Association of calcium and dairy product consumption with childhood obesity and the presence of a Brain Derived Neurotropic Factor-Antisense (BDNF-AS) polymorphism. Clin. Nutr. (2018).

33. Harrel, F.E. Jr. Regression modeling strategies. With applications to linear models, logistic and ordinal regression, and survival analysis (2nd ed.) (Springer Series in Statistics, New York, USA, 2015).

34. Lawless, J. F. & Singhal, K. Efficient screening of non-normal regression models. Biometrics. 34, 318–327 (1978).

35. Akaike, H. A new look at the statistical model identification. IEEE Trans. Autom. Control. 19, 716–723 (1974).

36. Lamiquiz-Moneo, I. et al. Genetic predictors of weight loss in overweight and obese subjects. Sci. Rep. 9, 10770 (2019).

37. Livingstone, K.M. et al. Fat mass- and obesity-associated genotype, dietary intakes and anthropometric measures in European adults: The Food4Me study. Br. J. Nutr. 115, 440–448 (2016).

38. Ortega-Azorin, C. et al. Associations of the FTO rs9939609 and the MC4R rs17782313 polymorphisms with type 2 diabetes are modulated by diet, being higher when adherence to the Mediterranean diet pattern is low. Cardiovasc. Diabetol. 11, 137 (2012).

39. Crossman, A., Anne Sullivan, D. & Benin, M. The family environment and American adolescents’ risk of obesity as young adults. Soc. Sci. Med. 63, 2255–2267 (2006).

40. de Vinck-Baroody, O. et al. Overweight and obesity in a sample of children with autism spectrum disorder. Acad. Pediatr. 15, 396–404 (2015).

41. Wake, M., Hardy, P., Canterford, L., Sawyer, M. & Carlin, J.B. Overweight, obesity and girth of Australian preschoolers: prevalence and socio-economic correlates. Int. J. Obes. (Lond). 31, 1044–1051 (2007).

42. Yang, Y. et al. Combined effect of FTO and MC4R gene polymorphisms on obesity in children and adolescents in Northwest China: a case-control study. Asia Pac. J. Clin. Nutr. 28, 177–182 (2019).

43. Kuciene, R. & Dulskiene, V. Associations between body mass index, waist circumference, waist-to-height ratio, and high blood pressure among adolescents: a cross-sectional study. Sci. Rep. 9, 9493 (2019).

44. Scheepers, L.E. et al. The intestinal microbiota composition and weight development in children: the KOALA Birth Cohort Study. Int. J. Obes. (Lond). 39, 16–25 (2015).

45. Rampelli, S. et al. Pre-obese children’s dysbiotic gut microbiome and unhealthy diets may predict the development of obesity. Commun. Biol. 1, 222 (2018).

46. Bai, J. Hu, Y. & Bruner, D.W. Composition of gut microbiota and its association with body mass index and lifestyle factors in a cohort of 7-18 years old children from the American Gut Project. Pediatr. Obes. 14, e12480 (2019).

47. Nirmalkar, K. et al. Gut microbiota and endothelial dysfunction markers in obese Mexican children and adolescents. Nutrients. 10, 2009 (2018).

48. Karlsson, F.H. et al. Symptomatic atherosclerosis is associated with an altered gut metagenome. Nat. Commun. 3, 1245 (2012).

49. Tomás-Barberán, F.A., García-Villalba, R., González-Sarrías, A., Selma, M.V. & Espín, J.C. Ellagic acid metabolism by human gut microbiota: consistent observation of three urolithin phenotypes in intervention trials, independent of food source, age, and health status. J. Agric. Food Chem. 62, 6535–6538 (2014).

50. Cortés-Martín, A. et al. Urolithin metabotypes can anticipate the different restoration of the gut microbiota and anthropometric profiles during the first year postpartum. Nutrients. 11, 2079 (2019).

51. Cecil, J.E., Tavendale, R., Watt, P., Hetherington, M.M. & Palmer, C.N. An obesity-associated FTO gene variant and increased energy intake in children. N. Engl. J. Med. 359, 2558–2566 (2008).

52. Almeida, S.M. et al. Association between LEPR, FTO, MC4R, and PPARG-2 polymorphisms with obesity traits and metabolic phenotypes in school-aged children. Endocrine. 60, 466–478 (2018).

53. Sandilands, A.J. & O’Shaughnessy, K.M. The functional significance of genetic variation within the beta-adrenoceptor. Br. J. Clin. Pharmacol. 60, 235–43 (2005).

54. Dionne, I.J. et al. Association between obesity and a polymorphism in the α-1 adrenergic receptor gene (Gly389Arg ADRB1) in Caucasian women. Int. J. Obesity. 26, 633–639 (2002).

55. Linné, Y., Dahlman, I. & Hoffstedt, J. β1-adrenoceptor gene polymorphism predicts long-term changes in body weight. Int. J. Obes. (Lond). 29, 458–462 (2005).

56. Aradillas-García, C. et al. Obesity is associated with the Arg389Gly ADRB1 but not with the Trp64Arg ADRB3 polymorphism in children from San Luis Potosí and León, México. J. Biomed. Res. 31, 40–46 (2016).

57. Lafontan, M. & Langin, D. Lipolysis and lipid mobilization in human adipose tissue. Progress Lipid Res. 48, 275–297 (2009).

58. Vargovic, P. et al. Adipocytes as a new source of catecholamine production. FEBS Lett. 585, 2279–84 (2011).

59. Lee, Z.S. et al. Urinary epinephrine and norepinephrine interrelations with obesity, insulin, and the metabolic syndrome in Hong Kong Chinese. Metabolism. 50, 135–43 (2001).

